# Development of a Rule-Based Natural Language Processing Algorithm to Extract Sleep Information in Pediatric Primary Care Patients with a Sleep Diagnosis

**DOI:** 10.1101/2025.05.31.25328640

**Authors:** Joseph W. Sirrianni, Ariana Calloway, Syed-Amad Hussain, Deena Chisolm, Kelly Kelleher, Azizi Seixas, Hongfang Liu, Christopher Bartlett, Mattina A. Davenport

## Abstract

**Introduction:** Retrospective analysis of sleep health among pediatric patients can enable important care and condition related discoveries. Often, sleep health is only encoded in a patient’s structured data after formal diagnosis. However, their unstructured clinical text often contains many detailed sleep health mentions prior to diagnosis. These mentions are numerous and cannot reasonably be identified manually, thus computer assisted tools must be developed. We present a novel, low-resource sleep vocabulary that can be applied to identify notes containing sleep mentions automatically.

**Methods:** Using a combination of existing sleep ontologies, interviews with clinicians, and examination of clinical note narratives, we develop a novel vocabulary of sleep health terms and phrases that cover both technical terms, abbreviations, and colloquial keywords used in describing sleep health. We compare our vocabulary against a set of manually annotated clinical notes to determine the effectiveness of our vocabulary for identifying notes with sleep health mentions.

**Results:** Our vocabulary was able to correctly identify clinical notes with sleep health mentions with a precision of 0.838 and recall of 0.869.

**Conclusion:** Our vocabulary showed excellent performance for identifying sleep health mentions at the clinical note level. The vocabulary was not able to accurately identify the specific text spans containing the mentions, which likely would require a more high-resource model. Thus, our low-resource vocabulary, which can be deployed in almost any compute environment, can serve as an identifying first pass over clinical notes to identify which notes should be further processed by more advanced models or manual review to identifying sleep health mentions.

## Introduction

It is established that sleep is a critical factor in adolescent development [Williamson et al. 2020, El-Shiekh et al. 2022], however many youths across the United States report getting inadequate sleep [Jesmin and Amin 2025]. While hospitals and clinics often gather some sleep related information about their patients during an encounter, the system broadly still strongly relies on either referrals to sleep care services or explicit patient/parent mention to identify inadequate sleep [Rubens et al. 2016, Williamson et al. 2016, Erichsen et al. 2012, Min et al. 2023]. This reliance on referrals and self-identification results in many patients being under-diagnosed for sleep related issues each year. Due to its significance in developmental health, it is imperative that sleep related issues are identified as early as possible at critical points in childhood development. Thus, there is much interest in the research community in developing methods to identify sleep related issues earlier in the identification process.

With the broad adoption of electronic health records (EHRs) in the United States, patient historical data, including their clinical notes, is easily accessible to providers for analysis. While the majority of EHR-based sleep related research has been conducted using structured fields, polysomnography data, and wearable technology [Perez-Pozuelo et al. 2020, Watson and Fernandez 2021, Bandyopadhyay and Goldstien 2023], more attention is being paid to clinical notes as a source of sleep related information [Davenport et al., 2024].

We have observed at our site that in many cases of patients with sleep-related diagnoses, mentions of their sleep related issues often appeared in their clinical notes prior to their first diagnosis. In some cases, the sleep-mentions appeared in patients’ notes before any indication of a sleep mention in their structured data. Based on this observation, it is apparent that the ability to accurately identify sleep mentions in written clinical notes will provide a powerful tool for improving sleep issue discoverability in patients and eventually lead to a decrease in underdiagnosis.

Identifying sleep mentions in clinical notes is a difficult task for two main reasons. First, from our initial exploration, we observe that sleep mentions occur in a variety of types of notes across several departments [Davenport et al. 2024]. There is no uniform sleep screening protocol across our hospital, so sleep mentions could be mentioned in almost any context. Second, sleep mentions often show up in clinical narrative sections of the notes (the section often hand written by the provider, as opposed to a table generated by the EHR vendor) and can be expressed using formal clinical terminology (e.g. “obstructive sleep apnea”), informal clinical terminologies like acronyms (e.g. “osa”), and causal terminology quoting a patient (e.g. “parent says they have trouble breathing while asleep.”) [Davenport et al. 2024]. Therefore, the ideal solution to identifying sleep mentions needs to 1) address the variability of language in sleep mentions and 2) be able to process a large quantity of notes, since we will not be able to filter easily by note type or department without missing mentions.

Natural Language Processing (NLP) tools and techniques that have been leveraged to extract information from clinical notes [Hossain et al. 2023]. Vocabulary-based keyword searches over text are extremely computationally efficient and can be processed over a large quantity of notes, but do not easily deal with the variability of language. More advanced methods, such as deep learning models or large language models (e.g. ChatGPT), which are the best at addressing the variability of language, require expensive specialized hardware, are computationally expensive, and take a long time to process a single note. Therefore, it would be very expensive (both computationally and monetarily) to run every note through such a model. Instead, it is significantly more efficient to do a first pass over the notes to filter out the notes that are most likely to contain a sleep mention, and then pass it to the more advanced model.

In this work, we propose a note filtering method using a vocabulary-based solution for identifying sleep mentions in clinical notes that can be applied at scale. We present a novel sleep-mention vocabulary set that expands upon our prior vocabulary set [Davenport et al. 2024] with the addition of both technical terms from several clinical ontologies, practical terms added from interviews with clinicians, and discovered terms by data mining our real world clinical notes at Nationwide Children’s Hospital.

We validate our solution against 300 clinical notes, manually annotated for sleep mentions, from across four departments over five years (2018-2023). We compared our vocabulary to Sivarajkumar et al.’s [2024] sleep vocabulary, the only other published sleep-specific vocabulary set to our knowledge. We find that our vocabulary is able to correctly recall 99.2% of all sleep mention containing notes, a 13.5% increase over Sivarajkumar et al’s [2024] vocabulary, while maintaining a similar level of precision.

Unlike complex NLP models, our proposed solution is a lightweight vocabulary set that can be applied to nearly all compute environments, including databases, and does not require dedicated hardware. Furthermore, our vocabulary is mutable, meaning sites can easily modify the vocabulary list to fit their site-specific needs.

Our contributions are as follows:

1. We present a novel vocabulary set named Davenport & Sirrianni Expanded (DSE) of sleep-related keywords which combine prior keyword banks, keywords from ontologies, keywords recommended by clinicians, and keywords discovered through data analysis on historic clinical notes.
2. We analyze the capability of the vocabulary set for identifying clinical notes with sleep mentions on real-world clinical notes from our site. We find that our vocabulary had a significant improvement in note recall while having negligible effect on precision, when compared to Sivarajkumar et al.’s (SEA) vocabulary set.
3. We discuss and propose a simple method for deploying this model as solution for identifying clinical notes with sleep mentions at scale. The method can easily scale up to 10,000s of notes in a repository. We discuss how this vocabulary set can be used as is for identifying sleep mentions or be the first step in a pipeline for identifying notes which can in turn be fed into precise, more computationally expensive, NLP models for specific identification.

## Background

Recently, there has been increased interest in utilizing NLP for sleep health related research [Hossain et al. 2023]. NLP has previously used for sleep related cohort identification and identification of sleep disorders in clinical notes [Mazzotti 2021]. Horner et al. [2022] developed a rule-based text mining algorithm for identifying sleep complaints in primary care notes, utilizing both keywords and regular expressions. They achieved a sensitivity of 53% and specificity of 91% on their holdout set.

While many studies utilize vocabulary sets that include sleep item, to our knowledge, only Sivarajkumar et al. [2024] has developed a vocabulary specifically for sleep. They utilized a keyword set to retrieve clinical notes and developed ruled-based, machine learning model-based, and Large Language model-based solutions to extract various sleep concepts from notes from Alzhiemer’s disease patients. They focused on seven concepts: snoring, napping, sleep problems, bad sleep quality, daytime sleepiness, night wakings, and sleep duration. For their rule-based approach, each concept developed a set of keywords and regular expressions using word embeddings. They found that their rule-based approach outperformed both machine-learning and the LLAMA 2 [Touvron et al. 2023] LLM with an F1-score spanning from 0.91-1.0 across their categories.

While Sivarajkumar et al.’s work focuses on adult Alzheimer’s patients, their vocabulary set is applicable to notes in a pediatric setting as well. While their work focused more on the methods for identifying sleep mentions within the text, which we leave to future work, than the identification of relevant clinical notes, which is our objective in this work, their vocabulary set is the only suitable comparison point to prior literature.

## Methods

### Clinical Note Data

Our cohort consists of youth between the ages of two and eighteen who received care between January 2018 to December 2023 from at least one of the following departments at our site: primary care, behavioral health, school-based, or healthy weight and nutrition. In total, 217,406 patients fit those criteria. Because sleep related issues are so often under-diagnosed, for this study, we opted to focus on patients with confirmed sleep issues. Thus, we pulled only patients that had a sleep diagnosis, which we define as a patient with at least one of the ICD-10 diagnosis listed in Table 1. Of the 217,406 patients, only 33,904 patients had a sleep diagnosis that fit our criteria. Furthermore, of the 33,904 patients with a sleep diagnosis, 2890 of them had no clinical notes from those four departments in our time period and therefore were omitted from the study. So, our final cohort consisted of 31,014 patients.

**TABLE 1:** List of ICD codes (ICD-10 and ICD-9) used to identify patients with sleep issues.

| **Sleep Disorder Category** | **Combined ICD Codes** |
| --- | --- |
| Circadian Rhythm disorders | G47.2, G47.20, G47.21, G47.22, G47.23, G47.24, G47.25, G47.26, G47.27, G47.29, 307.45, 327.3, 327.30, 327.31, 327.32, 327.33, 327.34, 327.35, 327.36, 327.37, 327.39 |
| Hypersomnia | F51.1, F51.11, F51.12, F51.13, F51.19, G47.1, G47.10, G47.11, G47.12, G47.13, G47.14, G47.19, G47.4, G47.41, G47.411, G47.419, G47.421, G47.429, 307.43, 307.44, 327.1, 327.10, 327.11, 327.12, 327.13, 327.14, 327.15, 327.19, 347, 347.0, 347.00, 347.01, 347.1, 347.10, 347.11, 780.53, 780.54 |
| Insomnia | A81.83, F51.01, F51.02, F51.03, F51.04, F51.05, F51.09, G47.0, G47.00, G47.01, G47.09, Z73.81, Z73.810, Z73.811, Z73.812, Z73.819, 046.72, 307.41, 307.42, 327.0, 327.00, 327.01, 327.02, 327.09, 780.51, 780.52, V69.5 |
| Parasomnia | F51.3, F51.4, F51.5, F98.0, G47.5, G47.50, G47.51, G47.52, G47.53, G47.54, G47.59, N39.44, 307.46, 307.47, 307.6, 327.4, 327.40, 327.42, 327.43, 327.44, 327.49, 780.56, 780.36 |
| Sleep Related movement disorders | G25.81, G47.6, G47.61, G47.62, G47.63, G47.69, 327.5, 327.51, 327.52, 327.53, 327.59, 333.94, 780.58 |
| Sleep related breathing disorders | E66.2, G47.3, G47.30, G47.31, G47.33, G47.34, G47.35, G47.36, G47.37, G47.39, P28.3, R06.83, 278.03, 327.2, 327.20, 327.21, 327.23, 327.24, 327.25, 327.26, 327.27, 327.29, 780.57 |
| Other sleep disorders | F51, F51.8, F51.9, G47, G47.8, G47.9, Y93.84, Z72.82, Z72.820, Z72.821, 307.4, 307.40, 307.48, 307.49, 327, 327.8, 780.5, 780.50, 780.55, 780.59, V69.4 |

We pulled clinical notes from patient EHRs into a comma separated value (CSV) files using SQL. The notes consist of progress notes and consults, span the years between 2018 and 2023, and come from four different departments: primary care, behavioral health, healthy weight, and school based. In total, there were 779,387 notes across all departments.

To determine which notes contained sleep-related mentions, we manually annotated a subset of 300 notes from the full note dataset, stratified by year and department. This ensured that we had a representative spread of notes that considers changes in department note taking style over time, which happens often at our site. This work was approved by the NCH IRB (STUDY00004027).

### Novel Keyword Bank Development

To increase the number of potential clinical notes containing sleep mentions, we develop a novel keyword bank, called Davenport & Sirrianni Expanded (DSE), which builds upon our prior vocabulary [Davenport, Sirrianni, and Chisolm 2024].

The DSE keyword bank was developed by expanding our prior vocabulary by consulting three different types of sources: clinical ontologies, consultations with clinicians, and text analysis of real-world clinical notes at our site. We made use of several clinical ontologies, including the Medical Subject Headings (MeSH) thesaurus [National Library of Medicine 2024a], UMLS [National Library of Medicine 2024b], SNOMED-CT [SNOMED International 2025], and LOINIC [Regenstrief Institute 2023], for technical terms related to sleep health. Additionally, we consulted with clinicians at our hospital about the types of terms they typically use in their documentation, along with any common abbreviations. Lastly, we utilized our in-house clinical note search engine, named DeepSuggest [Moosavinasab et al. 2021], to discover similar terms to those identified from the ontologies and clinicians based on word embedding similarities derived from our actual historic clinical notes.

During the keyword development, we began to separate the keywords in 37 distinct high-level categories. In practice, some keyword bank concepts showed up often in clinical note text in a context that was not related to sleep. For example, the Wheezing concept’s keyword would often show up for patients with asthma and didn’t have to do with sleep. We manually identified seven concepts that were not useful for identifying sleep mentions: Wheezing, Hyperactive, Dizziness, Daytime Mood, Tense, Inattention, and Nighttime Anxiety. While these concepts often show up in sleep mentions, they are not useful for identifying sleep mentions. Thus, we separated our high-level concepts into two Tiers. Tier 1 concepts were used in our note identification model and Tier 2 Concepts and do not include them in the model but were included in future analysis. The DSE keyword bank, associated high-level categories, and the category Tier assignments are shown in Table 2.

**TABLE 2:**
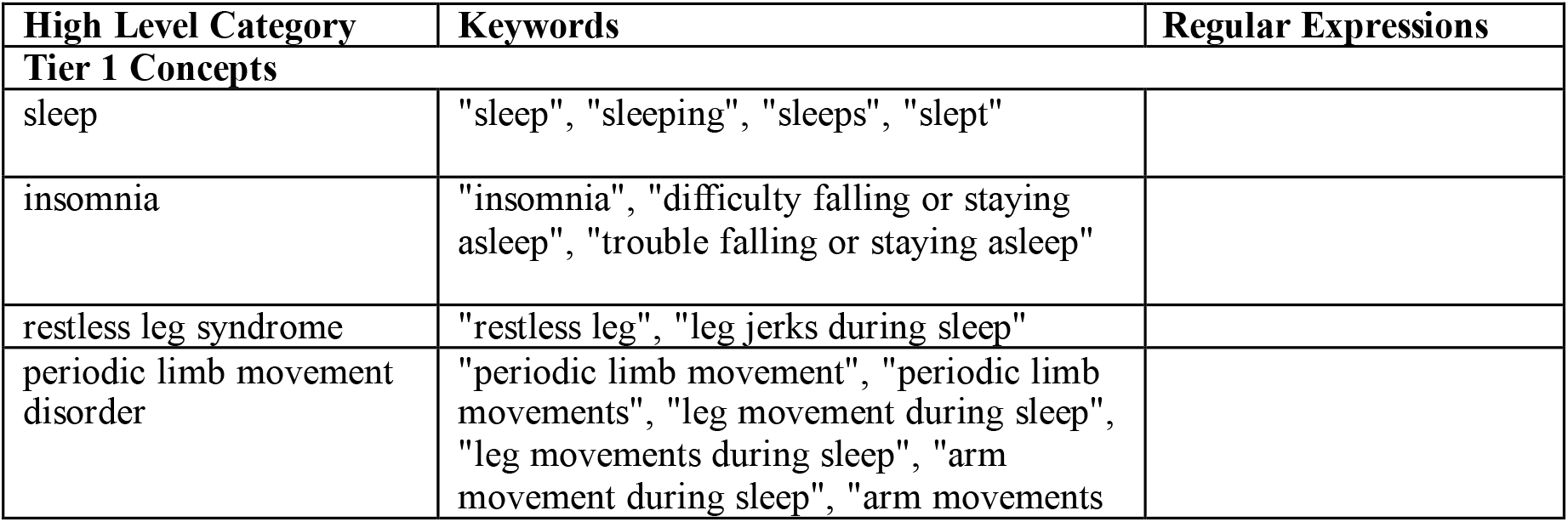
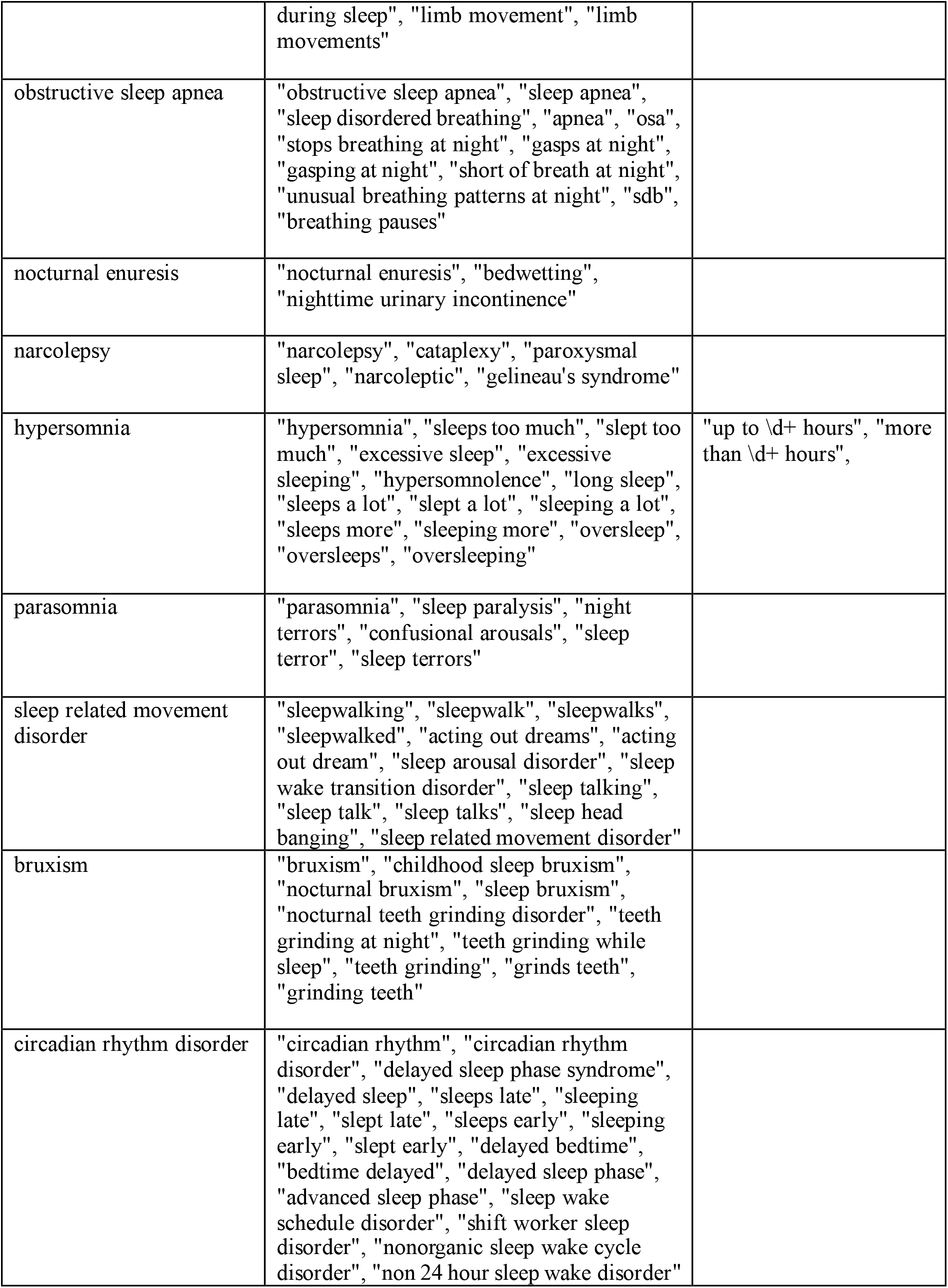
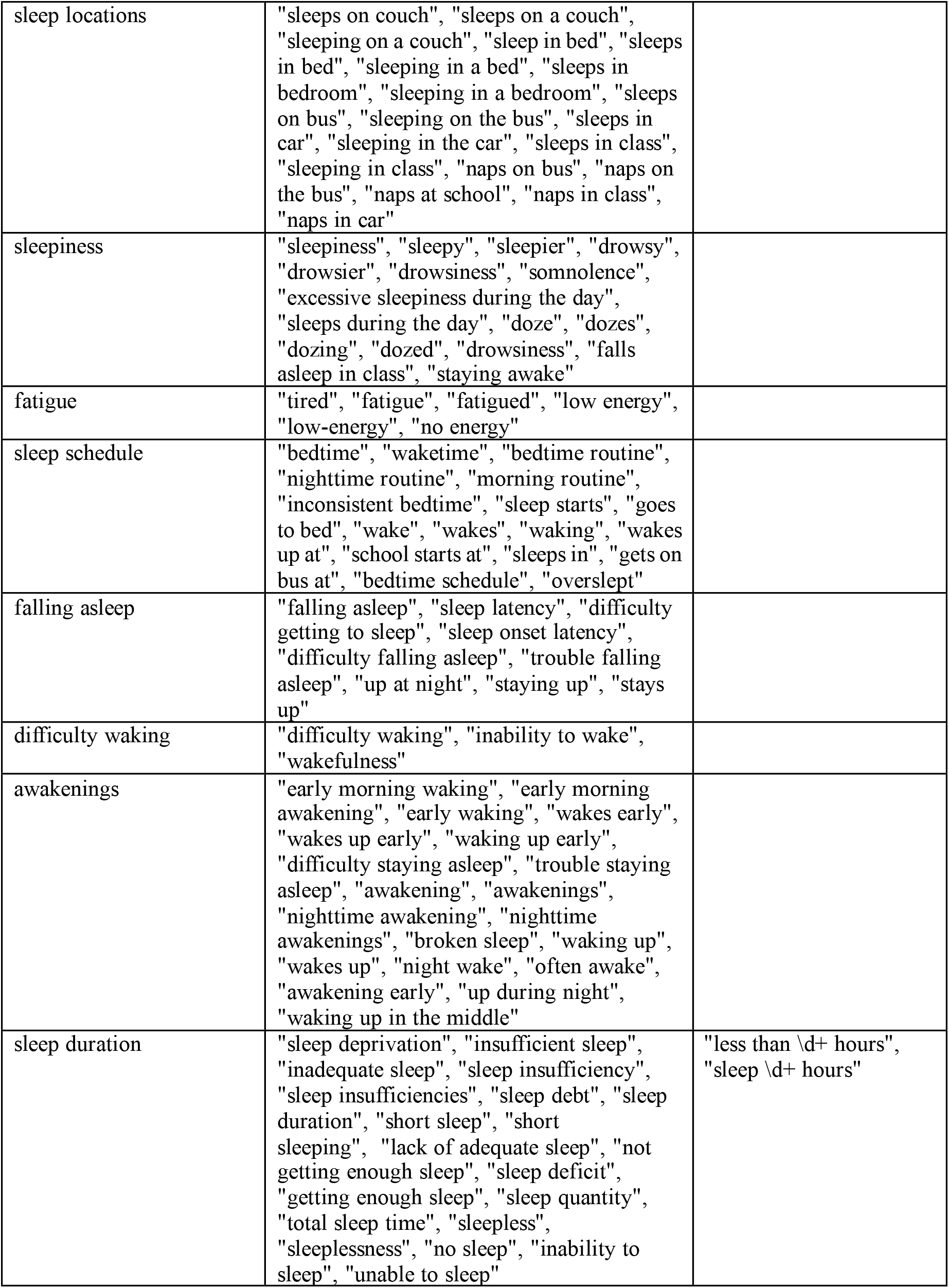
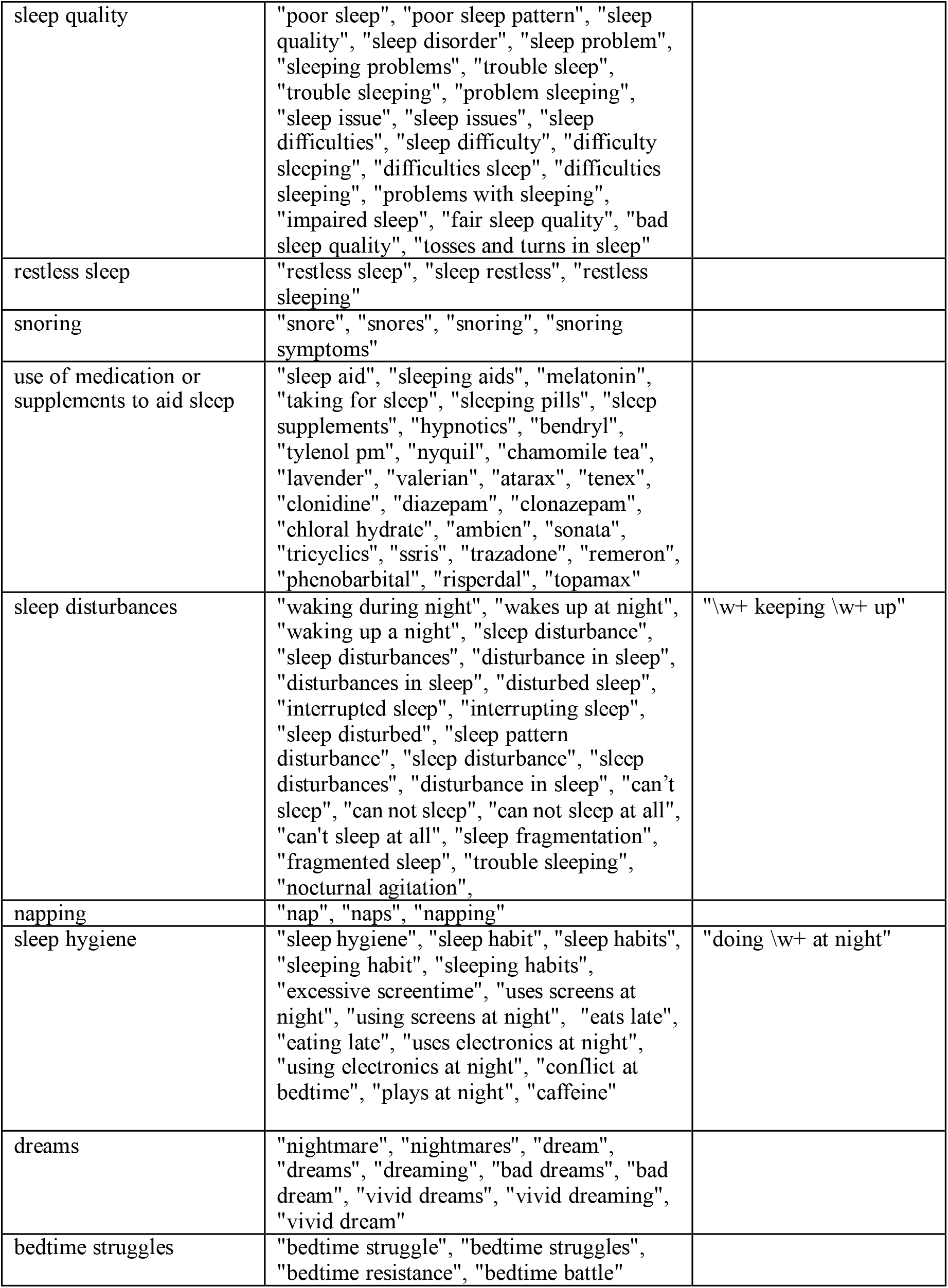
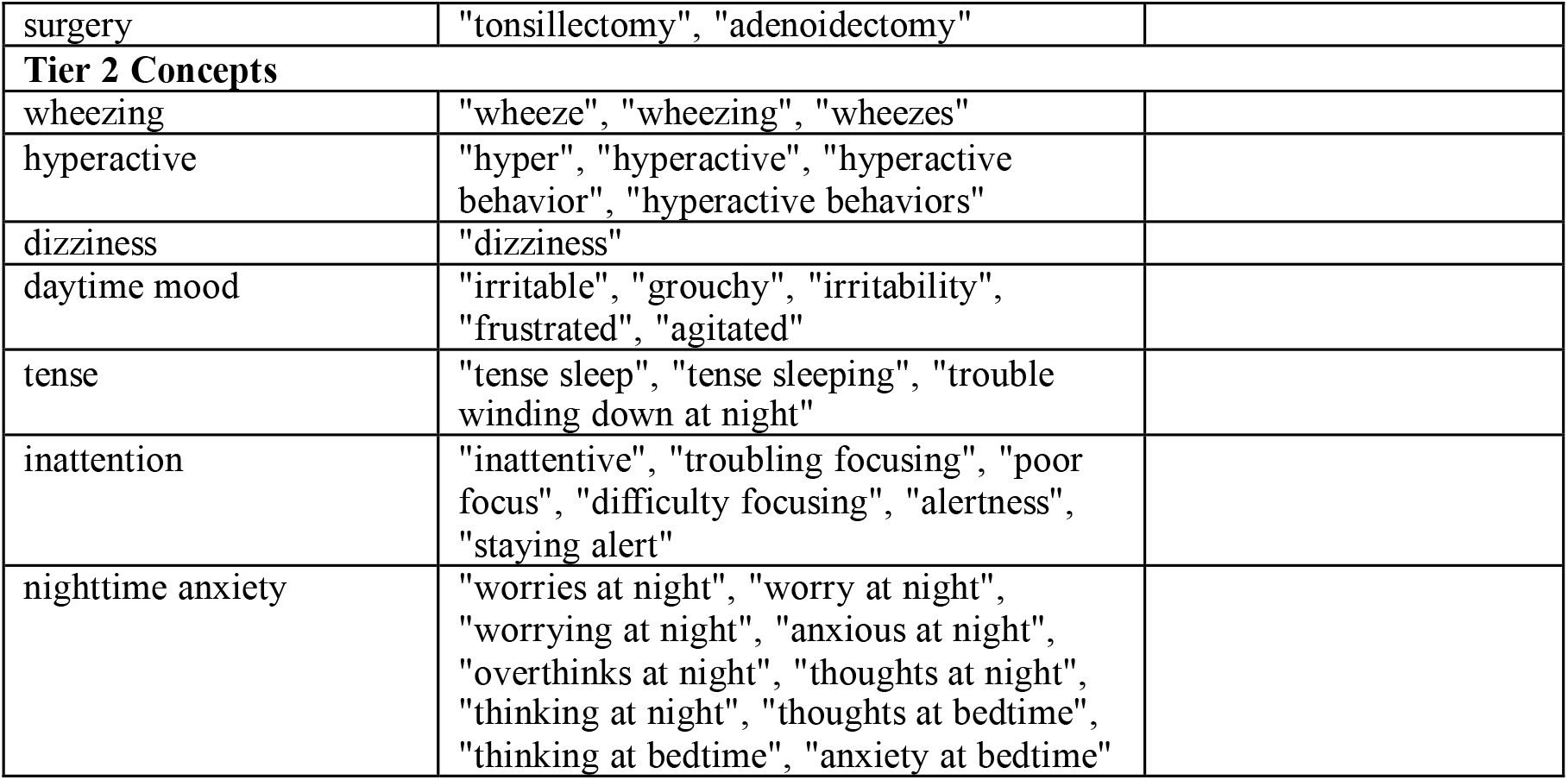
Davenport and Sirrianni Extended (DSE) keyword bank.

### Sleep Mention Annotation

Two annotators annotated the 300 notes for nine dimensions of sleep mentions: Sleep Behavior Dimension, Sleep Satisfaction Dimension, Alertness and Daytime Sleepiness Dimension, Sleep Timing Dimension, Sleep Efficiency Dimension, Sleep Duration Dimension, Sleep Medication Mentions, Sleep Disorder Mentions, and Sleep Intervention Mentions. The annotations were performed at the text span level, meaning that specific words and sentences were highlighted, using the tool MedTator [He et al. 2022]. Annotator disagreements were resolved by consensus between annotators.

While the annotations occurred at the text span level, for our evaluation we only focus on identifying notes if they contain any sleep mentions. Thus, for each note, we assigned an overall label to be either positive for sleep mentions if the note contains any tag and negative if it contains no tags.

## Modeling

### Keyword-Matching Model

We created a set of keyword models to identify if an input clinical note contains a sleep mention or not. Each model operates the same way and differ only by their vocabulary sets. The model follows a straightforward algorithm; the model is given a keyword bank to search against and a set of notes to classify. For each note, the model will search the note’s text for each keyword, key-phrase, or regular expression in the vocabulary. If *any* match is found in the note, the model will classify it as a positive and if no keywords are found it will be classified as a negative.

The model was developed in python. The keyword banks contained two types of keywords: text strings and regular expression patterns.

Text strings are exact keywords or phrases to be matched with no variability. For example, “apnea,” “obstructive sleep apnea,” and “OSA” are all text strings. The text strings are searched for in the clinical note’s text using FlashText library [Singh 2017], which implements a variation on the Aho-Corasick algorithm [Aho and Corasick 1975] for string searching, which is much faster and more efficient than regular expression searches or text scanning searches. The text strings are searched such that they are case-invariant (i.e. upper/lower case is ignored) and match entire words only (e.g. “osa” would not be matched to “prop**osa**l” as it is not the entire word).

Regular expression patterns are text strings that allow some variability within them. For example, the phrase “sleep X hours” has the X as a place holder for any number, so “sleep 8 hours”, “sleep 2 hours” and “sleep 12 hours” all should match the phrase. As such, these regular expression patterns must be encoded so they only match the desired type of strings (e.g. “sleep all hours” should not match on “sleep X hours”). While slower to run, regular expressions can match complex patterns that ensure that some placeholder text is a number, a word, or some combination of numbers and letters. These regex patterns defined in the vocabulary are matched using the standard regular expression package re in python.

### Keyword Banks

We utilized two keyword banks in our analysis, Sivarajkumar et al. (SEA) keyword bank [Sivarajkumar et al, 2024] and our Davenport & Sirrianni Expanded (DSE) keyword bank. In terms of the differences between DSE and SEA. Most of SEA’s 27 keywords are contained in DSE’s 359 total keywords. Only “hypopnea”, “nocturnal”, “rem”, and “nrem” are missing from DSE. Both “wheezing” and “dizziness” are included in DSE, but as tier 2 concepts and not used for note identification.

## Evaluation

We evaluate the predictions from two keyword prediction models, one using the MEA keyword bank and one using our DSE keyword bank. We compare these predictions to our ground truth annotations. Since these models are at the clinical note level, we consider a clinical note as a positive for a sleep mention if it contains at least one sleep mention tag, otherwise the note is false.

For each model, we generate a confusion matrix for a binary classification. Using the confusion matrix, we calculate precision, recall, and F1-scores to assess the performance of each model.

Precision is the ratio of the true positives (TP) over sum of the true positives and false positives (TP+FP). Precision values are on a scale from 0 to 1, where 1 indicates that model perfectly classifies positives without any false positive errors. Recall (also called sensitivity) is the ratio of true positives (TP) over the sum of the true positive and false negatives (TP+FN). Recall values are on a scale from 0 to 1, where 1 indicates that the model captures all of the positive cases with no false negatives. F1-score is the harmonic mean of precision and recall. F1-scores are on a scale from 0 to 1, where 1 indicates better overall model performance.

Additionally, we examined the prevalence of each individual keyword that appeared in the the true positives and false negative and calculate the keyword specific false positive rate. We report the top ten most prevalent keywords across the dataset and among the false positives. Lastly, we report how often the annotated tags contained keywords to gauge their recall.

## Results

Overall, there a total of 244 clinical notes containing sleep mentions and 56 note containing no sleep mentions. The confusion matrix for the DSE and SEA models are presented in Table 3a and 3b respectively. The DSE model had a total of two false negatives and 42 false positives, while SEA had 35 false negatives and 35 false positives.

**TABLE 3a:**
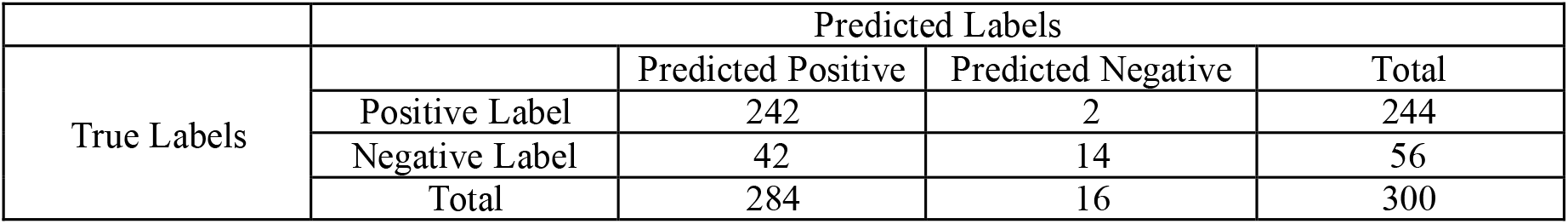
Confusion matrix for the DSE model.

**TABLE 3b:**
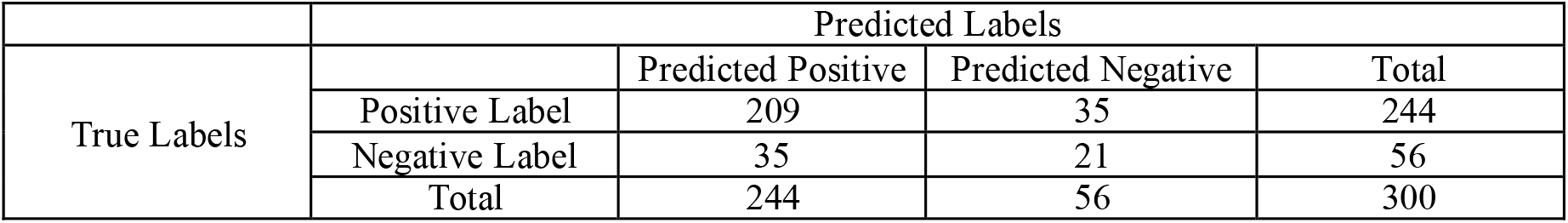
Confusion matrix for the SEA model.

The precision, recall, and F1 metrics are in Table 4. Overall, the DSE model had a much higher recall (0.992 vs 0.857) while having a comparable precision (0.852 vs 0.857). This difference results in a 0.06 difference in F1-score, driven by the increased recall.

**TABLE 4:** Precision, Recall, and F1-score metrics for the models.

|  | Precision | Recall | F1-score |
| --- | --- | --- | --- |
| MAE Model | 0.857 | 0.857 | 0.857 |
| DSE Model | 0.852 | 0.992 | 0.917 |

Table 5 shows the top 10 keywords from the DSE bank that appear across the dataset including the number of false positive notes and Table 6 shows the top 10 keyword from just the false positive notes. “Sleep” was the most commonly occurring keyword, appearing in 69.4% of all notes, however it was also the most common false positive occurring keyword, appearing in 71.4% of all false positives (False positive rate of 0.15). The other keywords had much lower false positive rates but occurred in fewer of the notes.

**TABLE 5:** Top 10 DSE (tier 1) keywords appearing across all notes.

| Rank | Keyword | Keyword Category | Total Note Appearances (%) | Total False Positive Appearances (False Positive Rate) |
| --- | --- | --- | --- | --- |
| 1 | Sleep | Sleep | 197 (69.4%) | 30 (0.15) |
| 2 | Bedtime | Sleep schedule | 88 (31.0%) | 4 (0.05) |
| 3 | Melatonin | Use of medication or supplements to aid sleep | 66 (23.2%) | 0 (0.00) |
| 4 | Alertness | Inattention | 49 (17.3%) | 0 (0.00) |
| 5 | Sleep disturbance | Sleep disturbances | 39 (13.7%) | 2 (0.05) |
| 6 | Sleeping | Sleep | 37 (13.0%) | 1 (0.03) |
| 7 | Fatigue | Fatigue | 35 (12.3%) | 1 (0.03) |
| 8 | Insomnia | Insomnia | 30 (10.6%) | 0 (0.00) |
| 9 | Tired | Fatigue | 28 (9.9%) | 2 (0.07) |
| 10 | Wake | Sleep schedule | 26 (9.2%) | 1 (0.04) |

**TABLE 6:** Top 10 DSE (tier 1) Keywords appearing in the false positive notes.

| Rank | Keyword | Total False Positive notes Keyword appears within. | Percentage of all False Positive Notes [N=42] |
| --- | --- | --- | --- |
| 1 | Sleep | 30 | 71.4% |
| 2 | bedtime | 4 | 9.5% |
| 3 | Morning routine | 3 | 7.1% |
| 4 | Adenoidectomy | 3 | 7.1% |
| 5 | apnea | 2 | 4.8% |
| 6 | tired | 2 | 4.8% |
| 7 | Sleep disturbance | 2 | 4.8% |
| 8 | alertness | 2 | 4.8% |
| 9 | Grinds teeth | 1 | 2.4% |
| 10 | tonsillectomy | 1 | 2.4% |

**TABLE 7:** Tagged Spans identified by DSE (tier 1) by dimension type.

| Tag Name | Total Tags | Identified Tags | Accuracy | Most Present Keyword (Total Tags) |
| --- | --- | --- | --- | --- |
| ALL TAGS | 1156 | 892 | 77.2% | Melatonin (101) |
| Alertness/ Day Time Sleepiness | 143 | 102 | 71.3% | Fatigue (41) |
| Sleep Satisfaction | 124 | 110 | 88.7% | Sleep (38) |
| Sleep Medications | 199 | 162 | 81.4% | Melatonin (101) |
| Sleep Disorder | 204 | 169 | 82.8% | Insomnia (44) |
| Sleep Intervention | 99 | 83 | 83.8% | Sleep (53) |
| Sleep Timing | 87 | 50 | 57.5% | Bedtime (13) |
| Sleep Behavior | 96 | 52 | 54.2% | Sleep (14) |
| Sleep Efficiency | 164 | 131 | 79.9% | Sleep disturbance (37) |
| Sleep Duration | 40 | 33 | 82.5% | Sleep (18) |

The total percentage of tagged spans in the notes that contain a keyword, broken down by tag type, are in Table 6. Across all tags, 77.2% contained at least one keyword from DSE. For each individual tag category, the keyword occurrence ranged from 88.7% (sleep satisfaction) to 54.2% (sleep behaviors).

## Discussion

Simple keyword/key-phrase models are easily implementable in most compute environments, including database queries, standard workstations, and mobile/edge devices. However, they lack context information to determine if a keyword mention is truly in the context of a sleep issue. This is made clear in our individual keyword breakdown where the most common word “sleep” appeared most often in the true positive *and* false positives. This shows the difficulty in identifying ideal keyword that have high recall and high precision. This trade-off is further exaggerated when looking at the actual annotated tagged spans. These results show that while our keywords do often show up in tags (77.2% of the time), they also show up outside the tags very often. More advanced NLP models can address the task of determining if a word is being used in a valid sleep mention context using technologies such as large language models. However, those tend to run very slowly compared to keyword search methods and require the use of highly specialized, expensive hardware, such as GPUs and TPUs. Thus, our focus was to develop a keyword-based model to be used to filter clinical notes such that only those that are likely to contain sleep mentions are identified by the model.

We found that our proposed keyword set, DSE, significantly out-performs the SEA vocabulary in terms of recall, while maintaining a similar level of precision on our notes. This is significant, as the intended use of the keyword model is to filter a large quantity of clinical notes quickly and efficiently to narrow down a subset.

Our development of this keyword bank included mining our site’s text data using word embedding similarity [Moonsavinasab et al. 2021] and interviews with providers on site, which likely gave us a large advantage over SEA, which is a generic keyword set developed for a cohort of adults. It seems likely that any site wishing to achieve high recall on a keyword model, would likely need to conduct a similar exploration of their site’s own data and note-taking habits to ensure the vocabulary is capturing all relevant sleep information.

This work has many limitations. As gestured at above, our vocabulary set was crafted for our specific site. We were unable to test it against any other sites so we cannot confirm our vocabulary works well in all circumstances. Second, as noted, some notes contain sleep vocabulary but not in the context of patient sleep issues. Our vocabulary-based filtering method cannot account for this and instead we rely on expert opinion as to which types of notes are likely to contain sleep mentions.

## Conclusion

In this work, we present a novel vocabulary set for pediatric sleep related sleep mentions developed from existing vocabulary sets and ontologies, terms identified through provider interviews, and terms identified through text mining. We use our vocabulary in an efficient note classification system. We compare our vocabulary against prior work and find that our vocabular set increases recall by 13.5% while maintaining precision. This work proposes the first step in a larger goal to automatically identify sleep mentions in clinical notes accurately, efficiently, and at scale. This identification will in turn eventually lead to earlier identification of pediatric sleep problems and improve outcomes.

## Data Availability

Data is not available.

## References

Aho, A. V., & Corasick, M. J. (1975). Efficient string matching: An aid to bibliographic search. Commun. ACM, 18(6), 333–340. 10.1145/360825.360855

Bandyopadhyay, A., & Goldstein, C. (2023). Clinical applications of artificial intelligence in sleep medicine: A sleep clinician’s perspective. Sleep and Breathing, 27(1), 39–55. 10.1007/s11325-022-02592-4

Davenport, M. A., Sirrianni, J. W., & Chisolm, D. J. (2024). Machine learning data sources in pediatric sleep research: Assessing racial/ethnic differences in electronic health record–based clinical notes prior to model training. Frontiers in Sleep, 3. 10.3389/frsle.2024.1271167

El-Sheikh, M., Gillis, B. T., Saini, E. K., Erath, S. A., & Buckhalt, J. A. (2022). Sleep and disparities in child and adolescent development. Child Development Perspectives, 16(4), 200–207. 10.1111/cdep.12465

Erichsen, D., Godoy, C., Gränse, F., Axelsson, J., Rubin, D., & Gozal, D. (2012). Screening for Sleep Disorders in Pediatric Primary Care: Are We There Yet? Clinical Pediatrics, 51(12), 1125–1129. 10.1177/0009922812464548

He, H., Fu, S., Wang, L., Liu, S., Wen, A., & Liu, H. (2022). MedTator: A serverless annotation tool for corpus development. Bioinformatics, 38(6), 1776–1778. 10.1093/bioinformatics/btab880

Horner, M., Abul-el-rub, N., Mays, M., & Mazzotti, D. (2022). 0610 Development of a Rule-Based Text Mining Algorithm to Identify Sleep Complaints in Primary Care Progress Notes. Sleep, 45(Supplement_1), A267–A268. 10.1093/sleep/zsac079.607

Hossain, E., Rana, R., Higgins, N., Soar, J., Barua, P. D., Pisani, A. R., & Turner, K. (2023). Natural Language Processing in Electronic Health Records in relation to healthcare decision-making: A systematic review. Computers in Biology and Medicine, 155, 106649. 10.1016/j.compbiomed.2023.106649

Regenstrief Institute (2023). LOINC®—Logical Observation Identifiers Names and Codes. https://loinc.org

SNOMED International (2025). SNOMED CT. https://www.snomed.org/snomed-ct

Irving, J., Patel, R., Oliver, D., Colling, C., Pritchard, M., Broadbent, M., Baldwin, H., Stahl, D., Stewart, R., & Fusar-Poli, P. (2021). Using Natural Language Processing on Electronic Health Records to Enhance Detection and Prediction of Psychosis Risk. Schizophrenia Bulletin, 47(2), 405–414. 10.1093/schbul/sbaa126

Jesmin, S. S., & Amin, I. (2025). Addressing the Sleep Deprivation Epidemic in Adolescents: Findings from the Youth Risk Behavior Survey 2021. American Journal of Health Education, 56(2), 142–151. 10.1080/19325037.2024.2366463

Mazzotti, D. R. (2021). Landscape of biomedical informatics standards and terminologies for clinical sleep medicine research: A systematic review. Sleep Medicine Reviews, 60, 101529. 10.1016/j.smrv.2021.101529

Min, J., Zhang, X., Griffis, H. M., Cielo, C. M., Tapia, I. E., & Williamson, A. A. (2023). Sociodemographic disparities and healthcare utilization in pediatric obstructive sleep apnea management. Sleep Medicine, 109, 211–218. 10.1016/j.sleep.2023.07.009

Moosavinasab, S., Sezgin, E., Sun, H., Hoffman, J., Huang, Y., & Lin, S. (2021). DeepSuggest: Using Neural Networks to Suggest Related Keywords for a Comprehensive Search of Clinical Notes. ACI Open, 05(1), e1–e12. 10.1055/s-0041-1729982

National Library of Medicine (U.S.). (2024a). Medical Subject Headings. https://meshb.nlm.nih.gov

National Library of Medicine (U.S.). (2024b). Unified Medical Language System (UMLS), 2024AA Release. https://www.nlm.nih.gov/research/umls

Perez-Pozuelo, I., Zhai, B., Palotti, J., Mall, R., Aupetit, M., Garcia-Gomez, J. M., Taheri, S., Guan, Y., & Fernandez-Luque, L. (2020). The future of sleep health: A data-driven revolution in sleep science and medicine. Npj Digital Medicine, 3(1), 1–15. 10.1038/s41746-020-0244-4

Rubens, S. L., Patrick, K. E., Williamson, A. A., Moore, M., & Mindell, J. A. (2016). Individual and socio-demographic factors related to presenting problem and diagnostic impressions at a pediatric sleep clinic. Sleep Medicine, 25, 67–72. 10.1016/j.sleep.2016.06.017

Singh, V. (2017). Replace or Retrieve Keywords In Documents at Scale (No. 1711.00046). arXiv. 10.48550/arXiv.1711.00046

Sivarajkumar, S., Tam, T. Y. C., Mohammad, H. A., Viggiano, S., Oniani, D., Visweswaran, S., & Wang, Y. (2024). Extraction of sleep information from clinical notes of Alzheimer’s disease patients using natural language processing. Journal of the American Medical Informatics Association, 31(10), 2217– 2227. 10.1093/jamia/ocae177

Touvron, H., Martin, L., Stone, K., Albert, P., Almahairi, A., Babaei, Y., Bashlykov, N., Batra, S., Bhargava, P., Bhosale, S., Bikel, D., Blecher, L., Ferrer, C. C., Chen, M., Cucurull, G., Esiobu, D., Fernandes, J., Fu, J., Fu, W., … Scialom, T. (2023). Llama 2: Open Foundation and Fine-Tuned Chat Models (No. 2307.09288). arXiv. 10.48550/arXiv.2307.09288

Watson, N. F., & Fernandez, C. R. (2021). Artificial intelligence and sleep: Advancing sleep medicine. Sleep Medicine Reviews, 59, 101512. 10.1016/j.smrv.2021.101512

Williamson, A. A., Meltzer, L. J., & Fiks, A. G. (2020). A Stimulus Package to Address the Pediatric Sleep Debt Crisis in the United States. JAMA Pediatrics, 174(2), 115–116. 10.1001/jamapediatrics.2019.4806

Williamson, A. A., Patrick, K. E., Rubens, S. L., Moore, M., & Mindell, J. A. (2016). Pediatric Sleep Disorders in an Outpatient Sleep Clinic: Clinical Presentation and Needs of Children With Neurodevelopmental Conditions. Clinical Practice in Pediatric Psychology, 4(2), 188–199. 10.1037/cpp0000135

